# Multi-modal data Integration for Metastatic Risk Prediction of AJCC Stage I/II Melanomas: Results from the Population-based Dutch Early-stage Melanoma Study (D-ESMEL)

**DOI:** 10.64898/2026.10.29.26366608

**Authors:** Jasper Ouwerkerk, Thamila Kerkour, Catherine Zhou, Antien Mooyaart, Andrew P. Stubbs, Marcel J.T. Reinders, Yunlei Li, Loes M. Hollestein

**Author notes:** Equal contributor (Yunlei Li, Loes M. Hollestein).

## Abstract

Early-stage melanoma accounts for 40-60% of melanoma-related deaths. Most prognostic modelling studies focus on unimodal approaches or a single multimodal integration strategy, overlooking alternative methods and biological processes underlying metastatic progression. We aimed to evaluate multi-modal data integration for predicting metastasis using the Dutch Early-Stage Melanoma (D-ESMEL) study, the world’s largest multi-modal cohort of American Joint Committee on Cancer (AJCC) stage I/II melanoma.

We analysed clinicopathological, genomic, transcriptomic, and pathomics data from the D-ESMEL study. The discovery set included 221 case-control sets matched on follow-up time, age, sex, Breslow thickness, and ulceration. The validation cohort included 154 AJCC stage matched case-control sets based on a population of >5,500 stage I/II melanoma patients. We evaluated four integration strategies (early, intermediate, mixed, and late) using a repeated (n=5) nested 3-fold cross-validation in the discovery set. The best-performing models were evaluated in the validation cohort using the odds ratios (ORs) and a weighted C-index reflecting performance in the sampled population.

In the discovery set, intermediate integration of genomics and transcriptomics achieved the highest balanced accuracy (µ: 0·62; 95% CI: 0·57-0·66). Early and late integration of transcriptomics and pathomics, and the unimodal transcriptomics model showed similar performance (µ: 0·61; 95% CI: 0·58-0·65). In the validation cohort both early integration, late integration, and the unimodal transcriptomics model achieved the highest balanced accuracy of 0·60· When adjusted for clinicopathological risk factors early integration remained an independent prognostic factor for metastasis (OR 1·75; p<0·01), outperforming transcriptomics alone (OR 1·26; p<0·01). Early integration reached a weighted C-index of 0·83 outperforming AJCC stage (0·81 C-index). When combined the population-weighted C-index was significantly improved to 0·85 (p<0·01).

Early integration of transcriptomics and pathomics outperformed all unimodal models and improved prediction of distant metastasis independently of AJCC stage. Understanding the biological mechanisms underlying these predictions may further support clinical decision-making.

## Introduction

Most melanoma patients are diagnosed at an early-stage (American Joint Committee on Cancer (AJCC) stage I/II) with a good prognosis.^1^ However, most patients who die due to melanoma were initially diagnosed with early-stage melanoma.^2^ This highlights the need for improved risk stratification, as the current AJCC staging system, based on Breslow thickness and ulceration, does not adequately identify the small set of stage I/II patients who are at high risk of metastasis. Therefore, additional prognostic biomarkers are needed to identify such high-risk patients.

Many studies have highlighted the potential of multi-modal data integration to discover novel prognostic biomarkers. However, existing studies remain constrained by key methodological and clinical limitations. Crucially, prior studies have not evaluated the effect of utilising different combinations of data modalities or integration strategies. Obscuring whether predictive performance is driven primarily by the data modalities utilised or by the integration technique, as different integration strategies utilising the same modalities and vice versa may reveal different results.^3–10^ Clinically, previous studies utilising data integration to improve prognosis have focused predominantly on overall survival.^3–6^ Currently, no data integration study specifically investigated metastasis prognosis in early-stage melanoma. Moreover, no previous study has validated its findings using a large validation cohort representative of a real-world melanoma population leaving clinical relevance and generalisability of their findings unassessed.

To address these limitations, we designed the Dutch Early-Stage Melanoma (D-ESMEL) Study, the largest collection worldwide of primary AJCC stage I/II melanoma samples to study progression to distant metastasis. The D-ESMEL study is comprised of a discovery set (442 samples) and a population-based validation cohort (308 samples selected from 5,785 stage I/II patients), including complete clinical data, histopathological imaging data, transcriptomic, and genomic sequencing data.^11,12^

Using this study, we aim to construct a framework to explore a diverse range of integration strategies (including early, late, mixed, and intermediate) encompassing diverse machine learning (ML) models to explore various techniques for a comprehensive evaluation of data integration strategies versus unimodal models.^13^ This study aims to further our understanding of the underlying biological patterns between multiple modalities and whether those patterns can improve metastasis risk stratification in early-stage melanoma patients.

## Methods

### D-ESMEL Study

The D-ESMEL study was designed to identify novel prognostic markers for early-stage melanoma based on multi-modal data. In brief, the discovery set is a matched case-control study, including 221 case-control sets (442 samples). Here cases are patient who developed distant metastasis and controls are patients who do not develop distant metastasis within the follow-up time of the case. Case-control sets are matched on age, sex, Breslow thickness and ulceration The validation cohort is a nested case-control (NCC) study, including 154 case-control sets with AJCC substage-matched controls (IA/IB/IIA/IIB/IIC) derived from >5,500 stage I/II melanoma patients selected from the Netherlands Cancer Registry. The validation cohort is used for the independent assessment of the added value of the best integrated models in a population-based cohort compared to AJCC stage alone. Moreover, prognostic value of data integration models can be assessed with sufficient statistical power in both stage I and II separately and be adjusted for baseline risk. All controls were selected using incidence density sampling, requiring potential controls to be at risk and event-free at the time of the matching case’s event. This design ensures that the calculated observed odds ratios mathematically approximate the incidence rate ratios of the full cohort.^14^ A detailed explanation of the complete design of the D-ESMEL study has been published elsewhere.^11,12^.

### Integration framework

Multi-modal data collected from the D-ESMEL study included targeted DNA mutation panel data (genomics), bulk RNA-seq (transcriptomics), automatically extracted histopathological features from H&E slides (pathomics), and manual clinicopathological scoring data (**Figure 1a**). Modality-specific pre-processing was conducted and missing values were imputed. These steps are described in detail in the **Supplementary Methods S1** and all included features across modalities are described in **Supplementary Table S1**. Samples missing any of the modalities were excluded and incomplete case-control sets were discarded. Finally, all modalities were fused into a singular matrix in which each row is a patient with complete data on all modalities.

**Figure 1.**
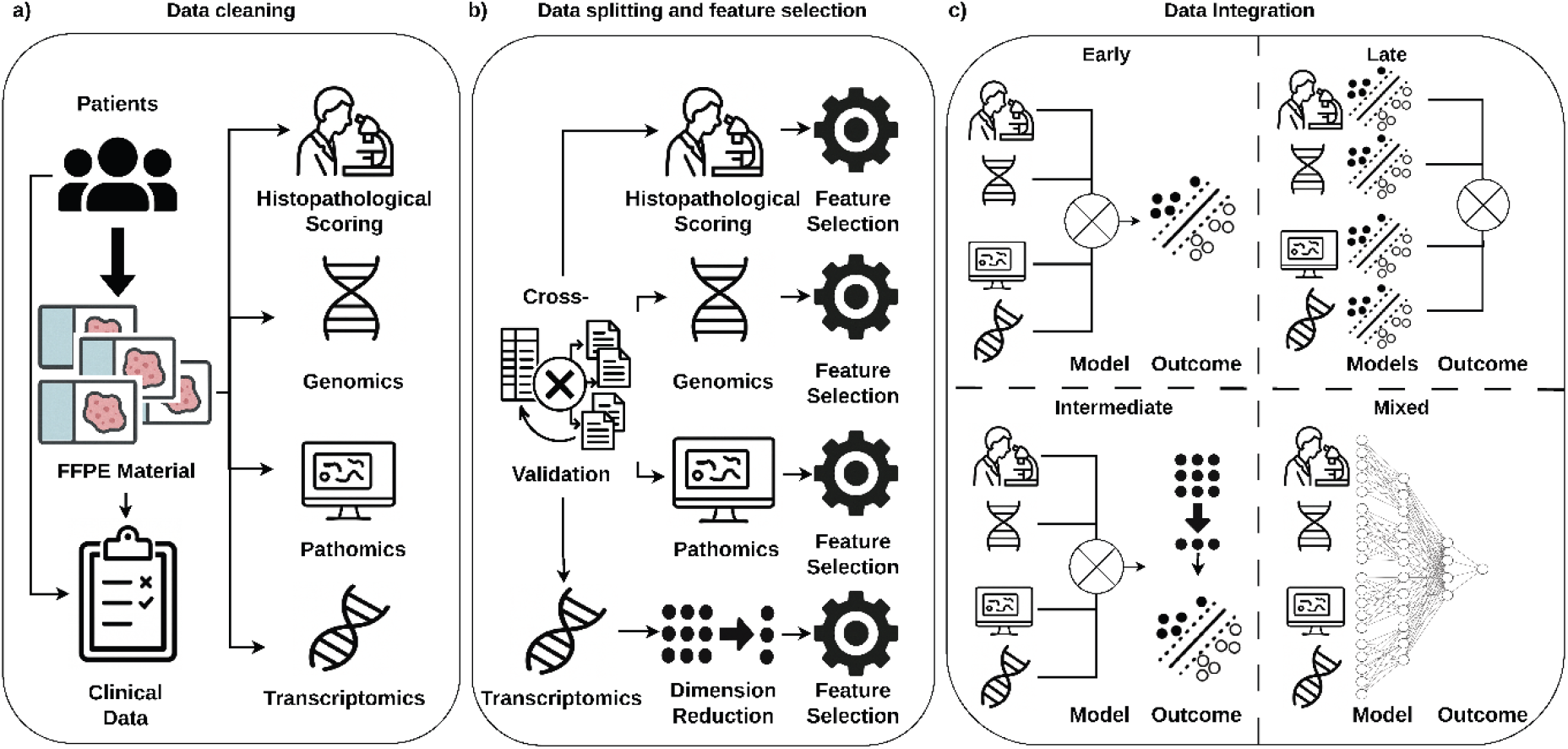
Flowchart of pre-processing the modalities and data integration. **a)** Data was generated from the patient’s formalin-fixed paraffin-embedded (FFPE) slides and case-control sets in the discovery set were divided into train, validation, and test folds using a five-time repeated 3-fold nested cross validation. **b)** Transcriptomic data was reduced in dimensions and for each modality the features were scaled and selected separately. **c)** Multiple data integration strategies were trained and evaluated. FFPE= formalin-fixed paraffin-embedded.

The remaining case-control sets in the discovery set were divided into training, validation, and test subsets using a repeated (n=5) 3-fold nested cross validation strategy (i.e. 3 outer folds and 3 inner folds). Dimension reduction was achieved through principal component analysis (PCA) and optionally followed by a Uniform Manifold Approximation and Projection (UMAP).^15^ Finally, a sequential feature selection step based on logistic regression was applied on each modality, testing both forward and backward selection on the training fold (**Figure 1b**).

Following the taxonomy proposed by Picard and colleagues^13^, four integration strategies were investigated: early, late, mixed, and intermediate (**Figure 1c**). Standard ML models were utilised for metastasis predictions including: logistic regression, support vector machine, random forest. More details on the data integration implementations are provided in the **Supplementary methods S1**.

#### Model Optimisation and Validation

The discovery set was used to optimise the workflow (**Figure 1**) and the validation cohort was used to evaluate the independent prediction performance. Balanced accuracy and area under the receiver operating characteristic (AUC) scores were calculated (**Supplementary Methods S1**).

All pre-processing algorithms and models were fitted on the training folds in the discovery set and the hyperparameters were optimised through a grid search across multiple dimensions of the workflow including input modalities, dimension reduction methods, feature selection methods, ML models, and data integration techniques, testing a total of 33,903 combinations (**Supplementary Table S2**). The best hyperparameters were selected based on the highest mean balanced accuracy in the 45 validation folds for each prediction approach, including multi-modal approaches (early/late/mixed/intermediate) and unimodal approaches (genomics/transcriptomics/pathomics/histopathological scoring). After hyperparameter tuning included data modalities in the validation cohort were based on the mean balanced accuracy in the 15 internal test folds and feasibility of clinical implementation. All models were then retrained on the complete discovery set and applied on all melanomas in the validation cohort for downstream analysis.

### Statistical Analysis

Univariable and multivariable conditional logistic regression models were applied to the validation cohort. Unweighted conditional logistic regression models were fitted using distant metastasis as the dependent variable to assess individual contributions of the unimodal models’ and the integrated multi-modal models’ predicted risk score besides the AJCC stage. The predicted risk score was rescaled to range from zero to one hundred. The independent variables in the multivariable model include the model’s rescaled predicted risk score, age, sex, location, and WHO subtype, since the controls were already matched on AJCC substage. Odds ratios (OR) of the best unimodal model and multi-modal model were assessed and the model with the largest significant OR was used for further statistical analysis.

Next, the effect of potential misclassification of stage III disease, due to the absence of a lymph node biopsy in some patients, was assessed by restricting the analysis to case-control sets with a negative sentinel lymph node (SLN) biopsy (SLNB) status and case-control sets with T1a, T1b, and T2a melanomas (n=160). These patients are either not advised to undergo SLNB or have an extremely low risk of SLN metastasis (<10%).^16^ Additionally, the statistical interaction between AJCC stage and the model’s predicted risk score was assessed using conditional logistic regression to test for equal performance across stages I and II.

A sample weighted Cox proportional hazards (CoxPH) model was fitted to assess the performance in the population-based validation cohort. The controls in the sampled population were weighted using a generalised linear model through the inverse sampling probability from the resized population cohort based on AJCC stage and follow-up time. The population cohort was resized based on the proportion of included case-control sets in the sampled population to maintain a comparable event rate in this source cohort.^17^ C-indices were compared using a log-likelihood test between the univariable CoxPH models using the integrated model’s risk prediction or AJCC stage and the multivariable CoxPH models combining both independent variables. To validate the integrated multi-modal model’s performance within a 5-year follow-up period, a 5-year time-dependent AUC analysis was performed using the log hazard ratios from the CoxPH models. A *p*-value < 0·05 was considered statistically significant.

### Explainability and Gene Set Enrichment Analysis

The best model’s predictions were interpreted through calculating Shapley Additive exPlanations (SHAP) values for each input variable in the validation cohort. SHAP values reflect the impact on the predicted outcome of a single input variable.^18^ The variables with the largest absolute SHAP values were further investigated. Variables created using PCA/UMAP were further analysed by calculating the contribution of each original variable towards a single latent feature of which the top 1% negative and positive contributors were selected. Involvement of biological pathways was analysed through an enrichment analysis of the Hallmark pathways.^19^ Genes involved within these pathways were clustered by their gene expression and clusters were checked for significant differences included modalities, clinical factors, and cell proportion calculated using EPIC.^20^

## Results

### Multi Modal Data Fusion

After fusing all data modalities and discarding any case-control sets with missing modalities, the discovery set consisted of 123 case-control sets (n=246) on which unimodal and integrated multi-modal models were developed. For the validation cohort 140 case-control sets (n=280) were included which are nested within a resized population-based cohort of 5,174 AJCC stage I/II patients. After fusing and pre-processing of all the modalities, a total of 39,996 features were available for model development (**Table 1**). Clinical characteristics of the discovery set and validation cohort are described in **Supplementary Table S3**.

**Table 1.** Overview of the number of samples and features across the different modalities included in the discovery set and validation cohort. Genomics data was not available for the validation cohort.

| Modality | Discovery | Validation cohort | Number of raw features | Number of included/pre-processed features |
| --- | --- | --- | --- | --- |
| *Histopathological scoring* | 442 | 308 | 49 | 26 |
| *Pathomics* | 366 | 308 | 25 | 10 |
| *Genomics* | 280 | - | 72 | 52 |
| *Transcriptomics* | 356 | 280 | 60,295 | 39,908 |
| *Fused* | 246 | 280 | 60,441 | 39,996 |

### Multi-modal Data Integration Outperforms Most Single Modality Models

Hyperparameter tuning in the discovery set resulted in an optimised model for each unimodal model and integration model (**Figure 2A**). In the validation folds, intermediate integration of genomic and transcriptomic data achieved the highest mean balanced accuracy (0·62). However, performance differed little from that of early integration, late integration, and the transcriptomic-only model (all around 0·61), indicating limited performance gains from increasingly complex integration strategies. Training balanced accuracy varies only slightly compared to the validation performance for each model, indicating no clear under- or overfitting.

**Figure 2.**
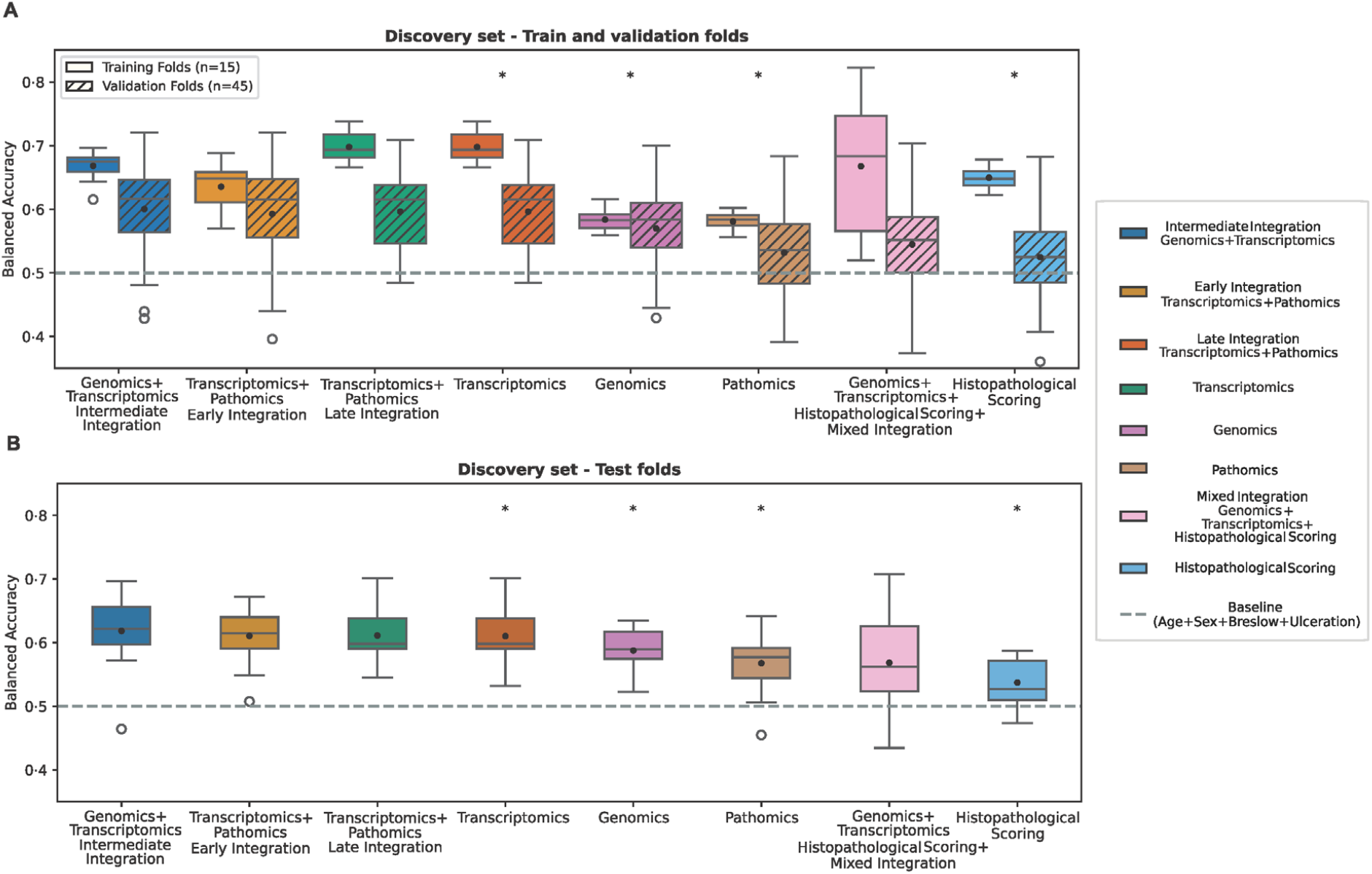
Best performing models based on the balanced accuracy across training, validation, and test folds in the discovery set. The dashed line at 0·50 represents the balanced accuracy using the matched clinical factors in the discovery set. **(a)** Balanced accuracy in the training and validation folds was comparable across intermediate, early, and late integration strategies, with the transcriptomic model also performing equally well. **(b)** Performance of the independent test folds in the discovery set revealing similar performance to the validation folds indicating generalisable results within the discovery set. Unimodal models are highlighted with *.

These optimised models were further evaluated for generalisability in the independent test folds and were compared to a baseline (0·50) which represents the effect of the matched clinical factors in the discovery set (**Figure 2B**). The balanced accuracy of intermediate integration, early integration, late integration, and the unimodal transcriptomics model were again similar (around 0·61) and not significantly different from each other (**Supplementary Table S4**). Overall, performance in the test folds and validation folds were similar indicating the models were generalisable within the discovery set. All models improved the performance compared to the matched clinical factors, but intermediate, early, and late integration significantly exceeded performance compared to mixed integration and the unimodal models (**Supplementary Table S4**). Finally, based on the minimal increase in performance of adding genomic data (mixed integration) and the high prognostic value of transcriptomics alone, we decided to include transcriptomics only, increasing the likelihood of future clinical implementation.

### Early integration improves metastatic risk prediction

The three best performing models (**Figure 2**) including transcriptomics data were selected to be retrained on the complete discovery set to utilise all available data to improve performance. Performance compared to AJCC stage and overall generalisability was assessed based on the AUC in the sampled population (**Figure 3a**) and the source population (**Figure 3b**) of the validation cohort. Additional performance metrics are described in **Supplementary Table S5**. In the sampled population the early integration model yielded stronger performance (AUC=0·66) compared to the late integration model and the unimodal transcriptomics model (AUC=0·64) (**Figure 3A**). These models outperformed AJCC stage (AUC=0·50) as its effect was matched out. In the source population AJCC stage the actual performance of AJCC stage is shown reaching an AUC of 0·81, which is better than both late integration and the unimodal transcriptomics model (AUC=0·79) (**Figure 3B**). However, early integration did reach a stronger performance (AUC=0·83) than AJCC stage.

**Figure 3.**
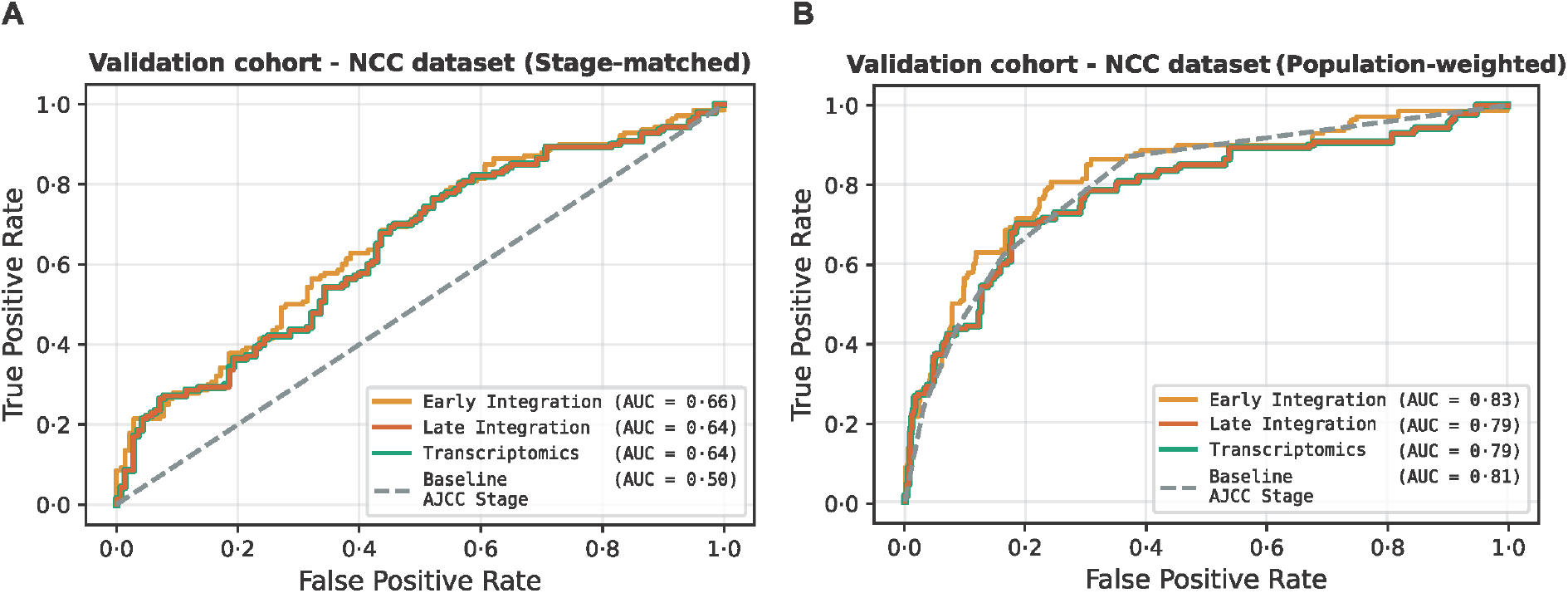
Performance in the validation cohort after retraining on the discovery set. a) Early integration reached strongest performance in the sampled population. b) Early integration showed a slightly better AUC than AJCC stage alone in the source population. AUC=Area under the receiver operating characteristic. AJCC=American Joint Committee on Cancer.

For the models ORs were calculated and adjusted for multiple clinical risk factors to assess the independence of the models’ predictions. Early integration was significantly associated (OR: 1.75; 95% CI: 1.50-2.04) with risk of metastasis (**Figure 4**). Late integration (OR: 1.48; 95% CI: 1.25-1.75) and the unimodal transcriptomics model (OR: 1.25; 95% CI: 1.14-1.37) also reached statistical significance, but showed a weaker association. The exact and adjusted ORs and CIs are described in **Supplementary Table S6**.

**Figure 4.**
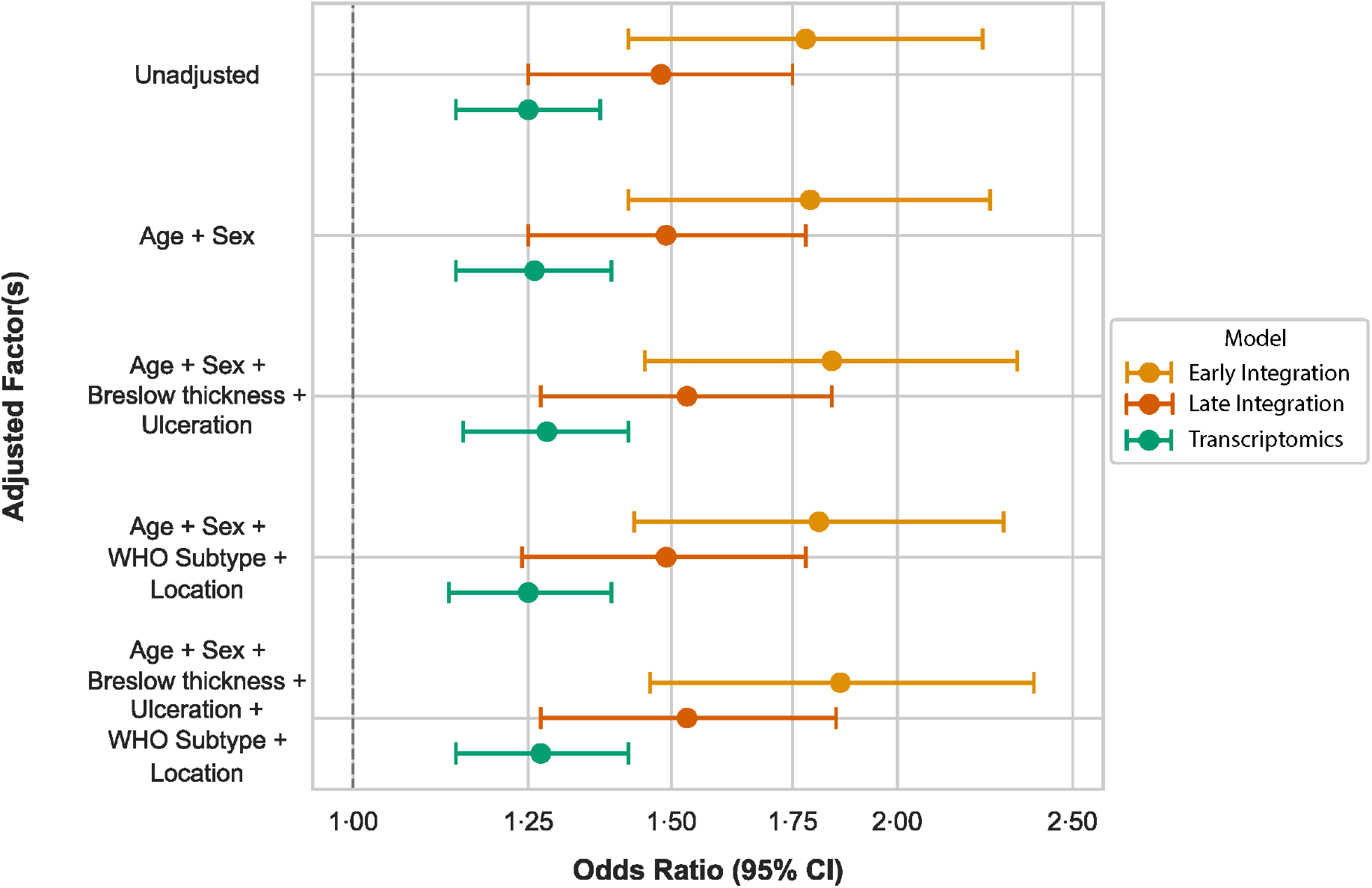
Odds ratios and 95% CIs of early integration, late integration, and the transcriptomics model in the sampled population. Early integration adds more prognostic value to AJCC stage than late integration or the transcriptomics model. AJCC= American Joint Committee on Cancer. CI= Confidence interval. OR= Odds ratios.

Early integration was further evaluated using a weighted CoxPH analysis. The early integration predictions showed strong prognostic power (C-index: 0·83; CI: 0·77-0·88) for metastatic risk and resulted in a significant (p-value< 0·01) improvement on top of AJCC stage alone (C-index: 0·81; CI: 0·75-0·86) reaching a C-index of 0·85 (CI: 0·79-0·89) (**Figure 5a**). The 5-year time-dependent AUC analysis were in alignment with the C-indices where the early integration model reached an AUC of 0·81, AJCC stage alone reached an AUC of 0·79, and the combination of the two achieved an AUC of 0·83 (**Figure 5B**).

**Figure 5.**
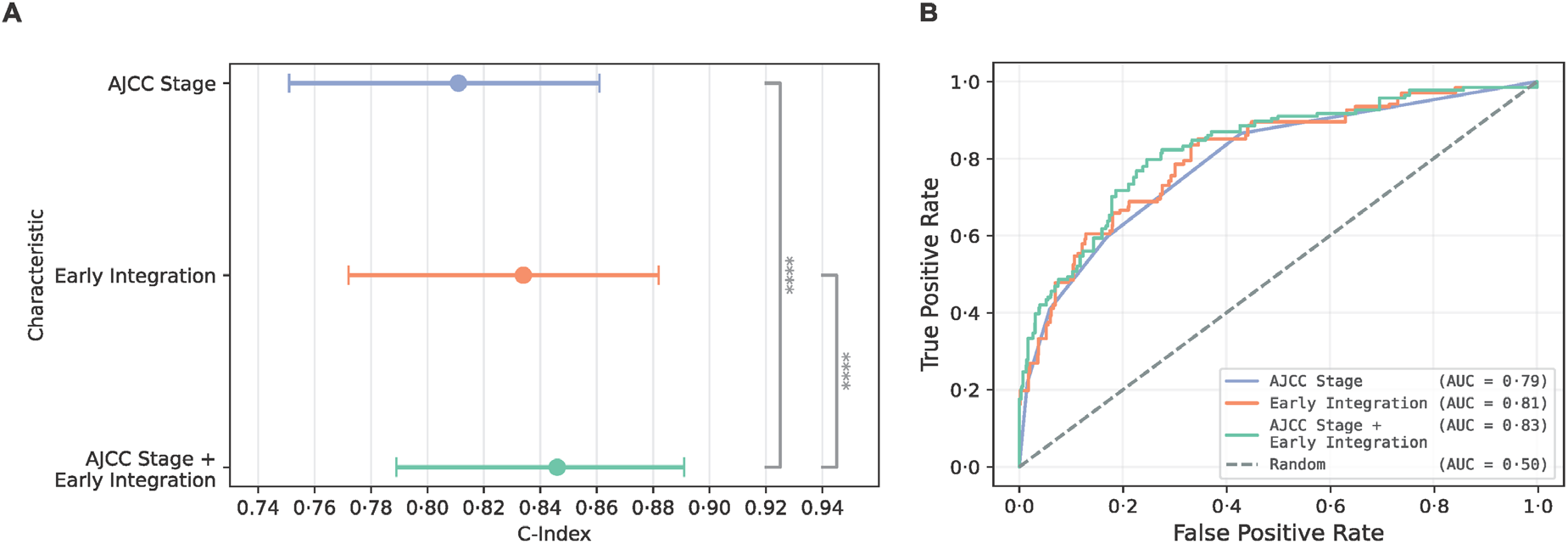
The prediction performances of the metastatic risk prediction from the early integration model, AJCC stage, and both variables combined in the validation cohort. **a)** CoxPH C-indices and standard errors. The integrated risk score (early integration) significantly adds to AJCC stage achieving a C-index of 0·85. **b)** The AUC curve show a similar improvement (AUC=0·83) within a 5-year follow-up timeframe when the metastatic prediction from the early integration is combined with AJCC stage. AJCC= American Joint Committee on Cancer. AUC= Area under the receiver operating characteristic. CoxPH= Cox proportional hazards.

### Sensitivity Analysis: Robustness of Integration Model

To assess the robustness of the early integration model, multiple sensitivity analyses were performed. No significant statistical interaction between AJCC stage and the early integration model’s metastatic risk predictions were found (p-value=0·09), revealing that the integrated model is equally valid for both stage I and II melanoma. In the subgroup of patients with a negative SLNB or with a very low risk of SLN metastasis (stage T1A/B, T2A), early integration’s metastatic risk predictions remained significant to predict distant metastasis (adjusted OR: 1.48; 95% CI: 1.12-1.97; p-value<0·01).

### Interpretation of early integrated features

The early integration approach utilises a Linear PCA to reduce the transcriptomic genes to 35 PCs, using backward feature selection, resulting in 32 transcriptomics PCs and 8 pathomics features being selected. These features were used in a logistic ridge regression model to predict metastasis. SHAP analysis revealed that the most contributing pathomics variables included mitotic rate (number of mitotic tumour cells per 30,000 tumour cells), tumour cell density, and the immune cell density in the tumour microenvironment (TME). Overall, the variables with the largest impact on the prediction of the early integration model (i.e. the highest absolute mean SHAP value) are three PCs constructed from transcriptomics data (**Figure 6a**). Higher values in PC1 and PC6 were associated with a higher risk for metastasis (i.e. a positive SHAP value) and vice versa. The inverse was true for PC0. The genes with the 1% largest positive and negative impact (i.e. loading value) towards these PCs (**Supplementary Table S7**) were found to be enriched in the epithelial mesenchymal transition (EMT) pathway, MYC signalling, and mTORC1 signalling (**Figure 6b**). Based on the expression of the genes enriched within these pathways, patients can clearly be separated into a highly proliferative (i.e. high cell proliferation and cell growth through MYC and mTORC1 signalling), a mesenchymal transition subgroup (i.e. high expression of the EMT pathway) subgroup, and a mixed/intermediate subgroup (similar expression of both proliferative and EMT pathways) (**Figure 6c**). PC6 was mainly made up of genes the EMT pathway (28/34 genes) and PC0 and PC1 mostly included genes from the proliferative pathways (59/71 genes) (**Supplementary Table S8**) Moreover, the mesenchymal subgroup had significantly more fibroblasts (**Figure 6e**) and increased immune cell density in the TME (**Figure 6d**). The proliferative group had the highest risk of developing distant metastasis and was significantly different (p-value<0·05) compared to the mesenchymal group which is associated with the lowest risk (**Figure 6f**). Risk of developing distant metastasis were also significantly different between mixed/intermediate (low) and the proliferative subgroup and between the mixed/intermediate (high) and mesenchymal subgroup (p-value < 0·05) (**Supplementary Table S9**).

**Figure 6.**
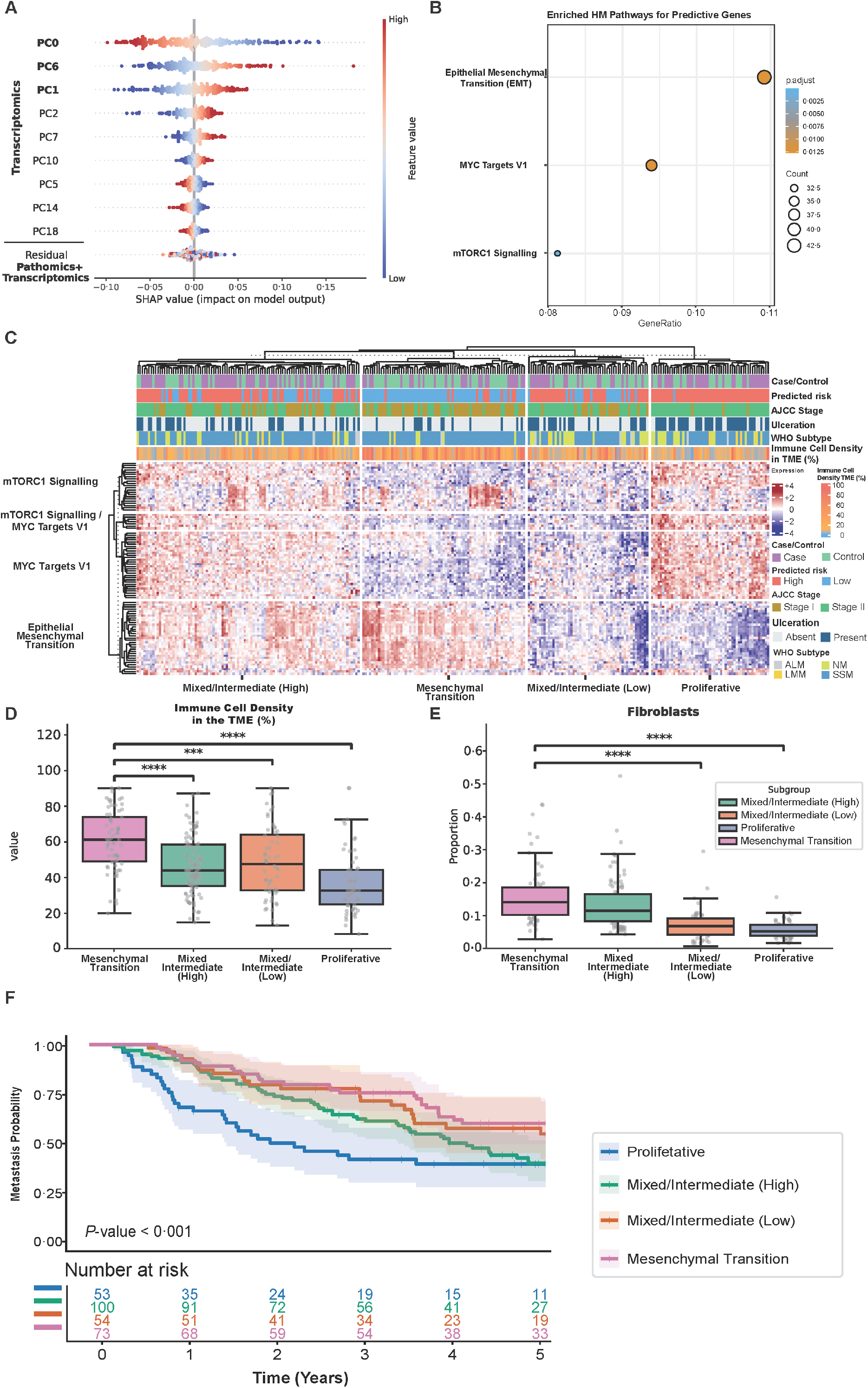
Feature interpretation of the early integration model predictive for prognosis of metastasis in the validation cohort. **a)** SHAP analysis of the top 10 most important features to predict metastatic risk using early integration. Larger values in PC0 are associated with a low risk and larger values in PC1 and PC6 with a higher risk for metastasis. **b)** The top 1% contributing genes were selected from PC0, PC1, and PC6, resulting in a set of 208 low risk and 208 high risk genes, which revealed enrichment of the MYC Targets V1, mTORC1 signalling, and the EMT pathway. **c)** Expression of the predictive genes within the enriched pathways, revealing four distinct subgroups (proliferative, mesenchymal transition, intermediate/mixed high, and intermediate/mixed low) differing in expression of the EMT and proliferative pathways. The intermediate subgroups did not show a single clear risk prediction. In addition, we observed that superficial spreading melanomas (SSM) and AJCC stage I patients were overrepresented in the mesenchymal group, and AJCC stage II were more common in the proliferative group. **d)** Immune cell density in the TME (%) is significantly higher in the mesenchymal subgroup compared to all other subgroups. **e)** The mesenchymal subgroup also had significantly more fibroblasts than the proliferative group. **f)** Kaplan-Meier curves of probability for developing distant metastasis in each subgroup over 5 years of follow-up. The proliferative subgroup’s had a significantly higher risk of developing metastasis compared to the mesenchymal subgroup, which had the lowest risk of developing distant metastasis (*p*-value<0·05). AJCC= American Joint Committee on Cancer. EMT= Epithelial mesenchymal transition. PC= Principle component. SHAP= Shapley Additive exPlanations. SSM= Superficial spread melanoma. TME= Tumour microenvironment.

## Discussion

This study presents a data integration framework for predicting distant metastasis in early-stage melanoma. The framework was developed on a matched case-control set to explore patterns among genomics, transcriptomics, and pathomics data that are prognostic for metastasis. Through rigorous study design from sample selection to comprehensive experiment setups utilising numerous ML and data integration techniques, we observed stronger performances in multi-modal models compared to unimodal models. Two multi-modal models integrating transcriptomic and pathomics data and one unimodal transcriptomic model, were validated for metastatic risk prediction in the population-based validation cohort 5,174 AJCC stage I/II melanoma patients using a nested case-control design. All models’ risk predictions were significantly associated with an increased risk for metastasis, but early integration demonstrated the strongest association. Importantly, our early integration model added independent and additional prognostic value on top of the AJCC staging in a population-based cohort, which could potentially further improve prediction of distant metastasis in early-stage melanoma.

Our multi-modal data integration framework utilised data of the D-ESMEL study. This allowed us to develop and validate our models on a large set of primary early-stage melanomas and assess performance independently and on top of AJCC stage. This highlights the uniqueness of our study as most previous data integration studies of melanoma focused on public data.^3–5,8,9^ Only a few previous studies utilised non-public data, but did rely on relatively small institutional cohorts (n=48, n=83, and n=283).^6,7,10^ This left the clinical relevance of their reported prognostic performance to real-world populations of patients with early-stage melanoma unverified. Moreover, none of these studies focused on prediction of distant metastasis in early-stage melanoma. Furthermore, previous studies might have overlooked potentially more effective integration strategies as all studies evaluated only one data integration strategy. Moreover, the contribution of each modality, performance of unimodal models, and different combinations of multi-modal inputs were rarely investigated, which limits the ability to determine if multi-modal models are truly more beneficial than unimodal models.^3–10^

These limitations are not unique to multi-modal integration studies of melanoma alone, as several multimodal studies across other cancer types relied on institutional cohorts and a single data integration method.^21–23^ Through our developed workflow, we optimised four different data integration techniques (early, late, mixed, and intermediate) and rigorously tested many broadly used ML models on all combinations of modalities, enabling a comprehensive evaluation of the prognostic benefit of each modality, unimodal models, and multi-modal data integration. By developing and validating this workflow using a nested case-control study, we safeguarded both the clinical relevance and reliability of our findings. In this study, we noted that multi-modal integration models generally achieved better overall performances, except for mixed integration which did not outperform unimodal models in the discovery set. This highlights that evaluating a single integration method approach can be insufficient.

Interpretation analyses of our early integration model revealed that increased proliferative signalling, cell growth, and lower immune cell density in the TME was associated with increased risk for metastasis. Moreover, we found a subgroup of melanomas defined by an increased expression of genes in the EMT pathway and increased immune cell density in the TME to be associated with a lower risk for metastasis. This is in line with other D-ESMEL studies which found increased immune cell density in the TME and specifically increased CD8+ in the TME to be associated with better outcomes.^24,25^ The enrichment of EMT signalling might be due to the significantly larger proportion of fibroblasts in the mesenchymal group compared to the proliferative subgroup, as cancer associated fibroblasts (CAFs) have been described to be a primary source of EMT signalling in the TME.^26^ The specific function and subtypes of the CAFs and how they affect metastatic risk specifically in this cohort remains unknown. In depth analyses of the state of the cell types present in the TME are needed to uncover which and how different cell types in early-stage melanoma interact and affects the risk of metastasis.

Our study is unique for its utilisation of a wide range of data modalities in a large well-defined cohort of early-stage melanomas to rigorously optimise and explore different data integration techniques. Firstly, we tuned the same hyperparameters for each pre-processing step and model for each modality, unimodal model, and multi-modal model using nested cross-validation to ensure generalisability and a fair assessment of each approach. Secondly, models were developed using data modalities across different biological layers to explore a wide range of biological variations. Thirdly, when developing the models, we used a matched case-control design based on known prognostic factors forcing the models to learn biological features independent of these already known prognostic clinicopathological risk factors. Finally, for model validation, we assessed the prognostic value in a population-based cohort using a NCC design. The validation of our findings in the independent population-based cohort further demonstrates that our models captured biologically relevant prognostic information.

Limitations of the D-ESMEL study design includes the use of aged FFPE material and not having a SLNB for all patients, which have been previously discussed.^11^ Extensive quality control of -omics data from aged FFPE material has been described previously. In this study we performed sensitivity analyses limited to SLN negative patients or patients not requiring a SLNB, to address this issue and we observed no difference. In the discovery set, the intermediate integration model with both genomics and transcriptomics modalities did not perform significantly better than the early integration model. As it is unlikely that both modalities will be performed in a clinical prognostic model with such a small difference in prognostic performance, we continued with transcriptomics analyses only. The current lack of genomic data in the validation cohort limits population-based genomic subtype analyses (i.e. BRAF-mutant, RAS-mutant, NF1-mutant, triple wildtype). However, cluster analyses in the D-ESMEL discovery set, including both genomic and transcriptomic data revealed that genomic subtype was not associated with transcriptomic subtype. In addition, for future research questions, which require information about the genomic subtype, isolated DNA is available for all samples.^11^ Finally, for the interpretability of the transcriptomics modality it has to be taken into account that the complete resection was sequenced not only tumor material.

In conclusion, we developed a framework for optimising multi-modal integration models, demonstrating the importance of comparing integration methods, modality combinations, and hyperparameters. Proliferative signalling and epithelial-mesenchymal transition emerged as major drivers for prediction of distant metastasis in early-stage melanoma. Further investigation of interactions within the TME may reveal mechanisms of metastasis and prognostic markers which could improve clinical decision-making and patient outcomes.

## Acknowledgements

We would like to thank the Netherlands Cancer Registry (NCR) for providing the clinical data. PALGA, pathology labs, collaborators for generating the data.

## Data availability

The clinical data used in this study were obtained from the NCR and can be requested through the Netherlands Comprehensive Cancer Organization. Data availability information can be found for each modality (histopathological scoring^24^, pathomics^24^, genomics, and transcriptomics^27^) in their corresponding articles.

## Ethics statement

Ethical approval was obtained from the scientific committees of Erasmus MC (MEC-2018–1738), the Dutch Nationwide Pathology Databank, and the Netherlands Cancer Registry.

The use of leftover diagnostic tissue samples for scientific research is based on the ‘no objection’ principle, as outlined in the Code of Conduct for Health Research by the Committee on Regulation of Health Research.

