## Supplementary Methods for "Multi-modal data Integration for Metastatic Risk Prediction of AJCC Stage I/II Melanomas: Results from the Population-based Dutch Early-stage Melanoma Study (D-ESMEL)"

### Data modalities

Formalin-fixed, paraffin-embedded (FFPE) specimens of the primary melanomas of the discovery set and validation cohort were collected across multiple diagnostic laboratories in The Netherland. Slides were cut and processed at the Erasmus University Medical Centre (Rotterdam, The Netherlands). These FFPE slides were used to extract genomic, transcriptomic, and histopathological information. A complete description of the tumor block collection and processing is described in the D-ESMEL study design.^1^

#### Genomics

Tumor material of the primary melanomas in the discovery set was sequenced using targeted Ion Torrent^TM^ sequencing based on a custom gene panel for melanoma.^2^ A gene was considered mutated if it harbored at least one pathological mutation. Presence of a mutation or CNV is denoted as 1 and absence as 0. Not all samples have complete information on genomic mutations or CNV status due to degradation of the DNA during the processes of FFPE. Therefore, missing values due to low DNA quality have been imputed by setting the mutation/CNV status to -1. Samples with unknown status in all mutations/CNVs are discarded. Additional explanation of the DNA sequencing and mutation calling is provided in another study of the D-ESMEL cohort.^2^

#### Transcriptomics

RNA was isolated from the primary melanomas, sequenced using the Illumina NextSeq™ 550 system, and transcript counts were extracted by SkylineDx BV (Rotterdam, The Netherlands, reference to Catherine article). The raw transcript counts were batch-corrected on the library kit and sequencing run using Combat-Seq ^3^. The corrected transcript counts are normalized using the transcript per million (TPM) normalization and converted to HUGO gene log2(TPM+1) normalized counts. Additional explanation of the bulk RNA sequencing and gene quantification is provided in another study of the D-ESMEL cohort.^4^

#### Clinical Histopathological Scoring

FFPE slides were stained using hematoxylin and eosin (H&E) and digitalized using the Hamamatsu NanoZoomer scanner at 40x magnification (0.225µm/px). Clinicopathological scoring data of the H&E whole slide images (WSI) were manually scored by AM.^5^

#### Pathomics

The histology features of the H&E WSIs are represented by the pathomics modality, calculated using a deep learning segmentation model that we previously developed and validated.^5,6^ The segmentation model extracts the areas of the whole tissue, tumor microenvironment, hair follicles, sebaceous glands, epidermis, ulceration and count data of tumor cells, immune cells, and mitotic cells. Missing values (~2% of the data) were imputed using the Multivariate Imputation by Chained Equations (MICE) method.^7^ MICE produces multiple versions of the original data. The multiple tables are concatenated into one table by extracting the median of continuous features and the most occurring value for categorical features.

**Modality feature selection**

To reduce noise and overfitting, bulk RNA-seq genes with low expression variance (<0·05) calculated within the training folds were excluded. Additionally, genes not expressed in more than 20% of the samples in the training folds were also excluded. All modalities were then standardized using robust scaling. The transcriptomic data underwent dimension reduction to balance the number of features across modalities.

**Data Integration**

In early integration modalities are directly concatenated and a single ML model is trained to predict metastasis. Also, late integration was implemented in which a separate ML model was fitted for each data modality separately and the final predictions were combined by majority voting, averaging of predicted outcomes, or an ensemble logistic regression model. Intermediate integration first transforms all modality features into a simplified representation through PCA/UMAP. Representations are created through PCA or UMAP on the concatenated modalities after the unimodal feature selection. These representations are used to train ML models. Finally, mixed integration involves the simultaneous integration of multi-modal data without requiring a pre-transformation step. To this end, a multi-headed neural network was implemented which extracted features from each modality individually through a neural network of which the extracted features are concatenated into a shared latent space. This shared latent space is propagated to another neural network layer which predicts metastasis. For all the other integration methods

**Model Validation**

Sensitivity, specificity, balanced accuracy, and the AUC score were calculated to evaluate performance. Sensitivity and specificity represent the accuracy of identifying cases (true positives) and controls (true negatives) respectively. Balanced accuracy is the mean of these two metrics, which shows how balanced the model performs between cases and controls. The AUC represents the overall ability of the model to distinguish cases from controls utilizing the predicted risk (0-1).

**Software**

Enrichment analysis was performed using the clusterProfiler-4.10.1 package. Cell proportions were calculated using EPIC-1.1.7, and Kaplan-Meier curves were generated using suvminer-0.5.2 in R-4.5.^8^
